# Platelet-rich fibrin in third molar surgery. Systematic review and meta-analysis protocol

**DOI:** 10.1101/2020.05.17.20099549

**Authors:** Gaston A. Salas, Shuheng A. Lai, Francisca Verdugo-Paiva, Roberto A. Requena

## Abstract

**Objective:** The objective of this systematic review is to assess the effectiveness and safety of platelet-rich fibrin (PRF) in third molar surgery.

**Data sources:** A comprehensive search strategy is meant to be used in an attempt to identify all relevant randomized controlled trials (RCTs), ongoing investigation reported in specialty congresses and trials regardless of language or publication status (published, unpublished, in press and in progress). Searches will be conducted in the Cochrane Central Register of Controlled Trials (CENTRAL), PUBMED, Embase, Lilacs, the International Clinical Trials Registry Platform (ICTRP), ClinicalTrials.gov, US National Institutes of Health (NIH), grey literature and in specialized congresses and conferences.

**Eligibility criteria:** We will include randomized trials evaluating the effect of PRF on wound healing after third molar surgery. Two reviewers will independently screen each study for eligibility, data extraction and risk of bias assessment using Cochrane ‘risk of bias’ tool. We will pool the results using meta-analysis and will apply the GRADE system to assess the certainty of the evidence for each outcome.

**Ethics and dissemination:** As researchers will not access information that could lead to the identification of an individual participant, obtaining ethical approval was waived.

## Introduction

Third molars are completely developed usually between 17 and 26 years of age^1,2^.There are three common scenarios for third molar eruption: complete eruption reaching functionality, partial exposition of the crown or full inclusion. Prevalence of third molars impaction rounds 24%^3^ which can be due to obstruction by another structure or abnormal path of development^2^. Specifically, impacted ones have been linked with diverse conditions such as pericoronitis, root resorption, periodontal disease, caries, cysts and tumors^4^. Diagnosis is generally made by a combination of clinical and radiographic assessment and its main course of treatment is surgical removal^5^.

In the last stages of the surgical procedure for third molars, the socket is prepared for healing in several ways. Standard care calls for socket conditioning and copious irrigation with saline solution, and closure by first or second intention. Multiple techniques have been proposed to aid and enhance wound healing to avoid further complications in third molar surgery such as pain, discomfort, bleeding, dehiscence, trismus and others^5^.

Platelet-rich fibrin (PRF) is a blood derivative obtained after centrifugation, conceived in 2001 by Choukroun^6^, to accelerate the healing of soft and hard tissue. It has the advantage to be completely autologous, not requiring any biochemical modification prior to its use and only needing one centrifugation cycle. This translates into a fast, simple and highly manageable biomaterial considering its adaptability to the receptor site^7,8^.

Although the use of PRF has been widely studied, the latest systematic reviews on the subject have several limitations regarding their results. Those reviews included moderate to high risk of bias RCTs with a high heterogeneity among them. On the other hand, some systematic reviews combined results from RCTs and retrospective studies decreasing its accuracy^9^ and others did not perform meta-analysis at all^10^. Recently published RCT may add information on whether the intervention modifies the outcomes assessed for the reasons mentioned above is expected then to find newer and higher quality RCTs that may deliver stronger conclusions supporting the use of PRF in third molar surgery.

The objective of this systematic review is to assess the effectiveness and safety of platelet-rich fibrin in third molar surgery compared to standard closure of wound with no material for enhancing healing.

## Methods

### Types of studies

All randomized controlled trials, both parallel and split-mouth designs, will be included. Non-randomized and quasi randomised controlled trials will be excluded.

### Types of participants

We will include trials assessing participants meeting the following criteria: males and females of all ages, with partially or fully erupted, impacted or not, third molars. Participants with third molars suffering from pathological signs (cysts, tumors), ectopic localization or aberrant anatomy, concomitant systemic disease and history of alcohol, tobacco or drug abuse will be excluded.

### Types of interventions

We will include trials evaluating platelet-rich fibrin as the final step of surgery prior to wound closure. After tooth extraction and standard conditioning of the tooth socket, PRF (PRF, L-PRF, A-PRF or other sub-types) is placed into it, surgical flap is then repositioned and sutured seeking healing by primary or secondary intention, this will be compared to no treatment of the post-extraction tooth socket or placebo. Studies that allowed concomitant use of pain medication will be included only if co-interventions are identical in both groups.

### Types of outcome measures

We will not use outcomes as an exclusion criterion during the selection process. Any article meeting all the criteria except for the outcome criterion will be preliminarily included and evaluated in full text. We will consider the following outcomes as relevance:

Primary outcomes:

- Alveolar osteitis
- Postoperative pain

Secondary outcomes:

- Swelling (edema)
- Infection
- Reintervention
- Adverse events
- Restricted mouth opening (Trismus)
- Clinical attachment loss

Other outcomes

- Soft tissue healing
- Bone density
- Bleeding

### Search methods for identification of studies

A comprehensive search strategy is meant to be used to identify all relevant RCTs, regardless of language or publication status (published, unpublished, in press, and in progress).

### Electronic searches

We will search the Cochrane Central Register of Controlled Trials (CENTRAL), PUBMED, Embase, Lilacs, WHO, International Clinical Trials Registry Platform (WHO-ICTRP) (www.who.int/ictrp/) and ClinicalTrials.gov (clinicaltrials.gov/) databases without language or date restrictions. Grey literature searches will be conducted to identify studies not indexed in the databases listed above in OpenGrey (www.opengrey.eu/) and the National Institute for Health and Clinical Excellence (NICE) (www.nice.org.uk/). The search strategy is presented on supplementary file.

### Searching other resources

We will review the reference lists of all included studies and relevant systematic reviews for additional potentially-eligible primary studies, contact the authors of eligible studies, researchers with expertise relevant to the review topic, conduct cited reference searches in ISI Citation indexes via Web of Knowledge and search in journals specialized in the field of oral and maxillofacial surgery. Abstracts and oral presentations of specialty meetings and congress (International Association of Dental Research and International Association of Oral and Maxillofacial Surgery, among others) will be reviewed as well.

### Data collection and analysis

Review authors (G.S. and S.L.) will independently screen titles and abstracts of all articles obtained through the searches against the inclusion criteria. Full texts will be obtained for all articles that seem to fulfill inclusion criteria and review authors will independently assess them to decide on their inclusion. Duplicates will be identified by comparing authors of the reports, trial dates, trial durations, number of participants, details on the interventions and location and setting of the reported trials and will be removed using collaboratron software. When multiple reports on one or more trials were identified these reports. Disagreements will be solved through a third party. Articles retrieved from the screening and included in the review will be recorded in the RevMan software^11^. Excluded trials after full text revision and the primary reason for the decision will be listed.

We will document the selection process in sufficient detail to complete a PRISMA flow chart^12^ and a table of ‘Characteristics of excluded studies’ as recommended in Section 11.2.1 of the *Cochrane Handbook for Systematic Reviews of Intervention*^13^.

### Data extraction and management

Review authors will, independently, extract data using a standardized form and check for agreement before data entry into Review Manager 5^11^. We are determined to collate multiple reports of the same study, so that each study, rather than each report, will be the unit of interest in the review. For each study with more than one control or comparison group for the intervention, other than “intervention” we will extract only the results of the control group “with no intervention”. We will not double count data within a meta-analysis, if different types of PRF are to be found, they will be combined into one single intervention, posteriorly, upon results, utility of stratification by types of PRF will be assessed. The following details will be recorded for each trial in the data extraction form: study design: randomized controlled trial (e.g. parallel, split-mouth, cluster); country of origin; setting (e.g. primary or secondary care); number of centers; recruitment period; funding source; inclusion criteria; exclusion criteria; number of patients randomized / number of patients evaluated; sealants and control, interventions (e.g. conventional treatments, preventive measures, no intervention); mode of administration; duration of interventions; primary and secondary outcomes; time(s) the outcomes were measured; method of sample size calculation; duration of follow-up; whether groups were comparable at baseline; any co-interventions; and any other issues

### Assessment of risk of bias in included studies

Included studies will be assessed independently by two review authors using the revised Cochrane risk of bias tool (RoB 2)^14^. Any disagreements will be resolved by discussion or by a third party.

We will evaluate each trial for the following domains:

1. Sequence generation
2. Allocation concealment
3. Blinding of participants and personnel
4. Blinding of outcome assessment
5. Completeness of outcome data
6. Risk of selective outcome reporting
7. Risk of other bias

Each domain will be judged in terms of high, unclear or low risk of bias, as described in Chapter 8 of the Cochrane Handbook^13^. These domains will inform an overall risk of bias, defined as follows:

- Low risk of bias: all domains are judged to be at low risk of bias.
- Unclear risk of bias: one or more domains are judged to be at unclear risk of bias.
- High risk of bias: one or more domains are judged to be at high risk of bias.

The assessments for each included study will be reported using RevMan 5^11^.

### Measures of treatment effect

For dichotomous outcomes, we will express the estimate of treatment effect of an intervention as risk ratios (RR) or odds ratios (OR) along with 95% confidence intervals (CI).

For continuous outcomes, we will use the mean difference and standard deviation (SD) to summarize the data using a 95% CI. Whenever continuous outcomes are measured using different scales, the treatment effect will be expressed as a standardized mean difference (SMD) with 95% CI. In cases where the minimally important difference (MID) is known, we will present continuous outcomes as MID units or inform the results as the difference in the proportion of patients achieving a minimal important effect between intervention and control.

If it is necessary, data from parallel-group and split-mouth trials will be combined using the procedure described by Lesaffre et al.^15^ and Elbourne et al.^16^. When needed, a paired analysis will be approximated by imputing the pooled standard deviation (SD) from the SDs of the 2 groups using a correlation coefficient of 0.5. All analyses used random-effects models.

### Dealing with missing data

We will contact study authors to obtain additional information whenever data appears to be missing or unclear. In order to undertake an intention-to-treat analysis, when possible, we will search data on the number of participants by the allocated treatment group, irrespective of compliance and whether or not the participant was later classified to be ineligible or otherwise excluded from treatment or follow-up.

### Assessment of heterogeneity

We will quantify the impact of statistical heterogeneity using the I^2^ statistic, which describes the percentage of total variation across studies that is due to heterogeneity rather than sampling error^13^. Where statistical heterogeneity is moderate, substantial, or high (I^2^ > 75%) or where there is clinical heterogeneity, we will investigate possible causes by exploring the impact of participants’ characteristics (e.g. wound etiology) or other variables. We will not pool studies which had high statistical heterogeneity (I^2^ > 75%) as this may produce misleading results.

### Assessment of reporting biases

We will assess reporting bias by comparing outcomes reported in the published report against the study protocol, whenever this could be obtained. If we could not obtain the protocol, we will compare outcomes listed in the methods section with those whose results were reported. If information is insufficient to judge the risk of bias, we will note this meta-analysis as having unclear risk of bias. If any meta-analysis includes a sufficient number of trials (more than 10), we will assess publication bias according to the recommendations on testing for funnel plot asymmetry, as described in Section 10.4 of the *Cochrane Handbook for Systematic Reviews of Interventions*^13^. If asymmetry is identified, we will examine possible causes or assess the asymmetry by using a table to list the outcomes reported by each study included in the review, to identify whether any studies did not report outcomes that had been reported by most studies.

### Data synthesis

We will only undertake meta-analysis for clinically homogeneous RCTs. We will undertake meta-analysis using the statistical package Review Manager 5, provided by Cochrane^11^. We will combine risk ratios for dichotomous data and mean differences for continuous data using the inverse variance method with the random-effects model. If combining outcome data is not possible due to differences in the reported outcomes, we will present a narrative summary.

### Subgroup analysis and investigation of heterogeneity

We will perform subgroup analysis according to:

- Type of PRF used (PRF, L-PRF or A-PRF)
- Position of the third molar (partially, included, fully erupted, impacted)
- Differences of equipment used to obtain PRF plugs, membranes and/or exudate.

In case we identify significant differences between subgroups (test for interaction <0.05) we will report the results of individual subgroups separately. The I^2^ index will be used to assess heterogeneity, which will be classified according to the following cut-off points: <40% low; 30-60% moderate; 50-90% substantial, 75-100% considerable. Any level of heterogeneity greater than 40% should be explained according to the covariates collected, by sensitivity analysis (elimination of a study), and by subgroup analysis and/or meta-regression. If heterogeneity cannot be explained, the results of the meta-analysis performed will not be offered.

### Sensitivity analysis

We will perform sensitivity analysis excluding studies with high risk of bias. In cases where the primary analysis effect estimates and the sensitivity analysis effect estimates significantly differ, we will either present the low risk of bias - adjusted sensitivity analysis estimates - or present the primary analysis estimates but downgrading the certainty of the evidence because of risk of bias.

### Summary of Findings

We will summarize the findings of the main intervention comparison by undertaking the following procedure to create a ‘Summary of Findings’ (SoF) table:

- Assessing the certainty of the evidence for each outcome using the GRADE approach
- Summarizing the findings for each outcome (quantitatively, where possible).
- completing the SoF table.
- Preparing bullet points that summarize the information in the SoF table in plain language.

Review authors will independently assess the certainty of the evidence (high, moderate, low, and very low) using the five GRADE considerations (study limitations, consistency of effect, imprecision, indirectness, and publication bias). We will use methods and recommendations described in Section 8.5 and Chapter 12 of the *Cochrane Handbook*^13^ and GRADEpro software^17^ to build SoF tables. We will resolve disagreements on certainty ratings by discussion, provide justification for decisions regarding the ratings using footnotes in the table, and make comments to aid readers’ understanding of the review where necessary. We will use plain language statements to report these findings.

## Data Availability

The authors declare that all the data supporting the findings of this study are available within the article and its supplementary information files.

## Ethics and dissemination

As researchers will not access information that could lead to the identification of an individual participant, obtaining ethical approval was waived.

## Authors’ contributions

G.S. conceived the protocol. G.S. and MF.V. drafted the manuscript, and all other authors contributed to it. The corresponding author is the guarantor and declares that all authors meet authorship criteria and that no other authors meeting the criteria have been omitted.

